# Relation of Quantitative Histologic and Radiologic Breast Tissue Composition Metrics with Invasive Breast Cancer Risk

**DOI:** 10.1101/2020.11.12.20230623

**Authors:** Mustapha Abubakar, Shaoqi Fan, Erin Aiello Bowles, Lea Widemann, Máire A. Duggan, Ruth M. Pfeiffer, Roni T. Falk, Scott Lawrence, Kathryn Richert-Boe, Andrew G. Glass, Teresa M. Kimes, Jonine D. Figueroa, Thomas E. Rohan, Gretchen L. Gierach

## Abstract

**Purpose:** Benign breast disease (BBD) is a strong breast cancer risk factor but identifying patients that might develop invasive breast cancer remains a challenge.

**Methods:** By applying machine-learning to digitized H&E-stained biopsies and computer-assisted thresholding to mammograms obtained circa BBD diagnosis, we generated quantitative tissue composition metrics and determined their association with future invasive breast cancer diagnosis. Archival breast biopsies and mammograms were obtained for women (18-86 years) in a case-control study, nested within a cohort of 15,395 BBD patients from Kaiser Permanente Northwest (1970-2012), followed through mid-2015. Cases (n=514) who developed incident invasive breast cancer and controls (n=514) were matched on BBD diagnosis age and plan membership duration.

**Results:** Increasing epithelial area on the BBD biopsy was associated with increasing breast cancer risk [Odds ratio(OR) 95% confidence interval(CI) _Q4 vs Q1_=1.85(1.13-3.04);P_trend_=0.02]. Conversely, increasing stroma was associated with decreased risk in non-proliferative, but not proliferative, BBD (P_heterogeneity_=0.002). Increasing epithelium-to-stroma proportion [OR(95%CI)_Q4 vs Q1_=2.06(1.28-3.33);P_trend_=0.002] and percent mammographic density (MBD) [OR(95%CI)_Q4 vs Q1_=2.20(1.20-4.03);P_trend_=0.01] were independently and strongly predictive of increased breast cancer risk. In combination, women with high epithelium-to-stroma proportion/high MBD had substantially higher risk than those with low epithelium-to-stroma proportion/low MBD [OR(95%CI)=2.27(1.27-4.06);P_trend_=0.005], particularly among women with non-proliferative [P_trend_=0.01] versus proliferative [P_trend_=0.33] BBD.

**Conclusion:** Among BBD patients, increasing epithelium-to-stroma proportion on BBD biopsies and percent MBD at BBD diagnosis were independently and jointly associated with increasing breast cancer risk. These findings were particularly striking for women with non-proliferative disease (comprising ∼70% of all BBD patients), for whom relevant predictive biomarkers are lacking.

## Introduction

In the U.S., >70% of 1.6 million annual breast biopsies are benign (1, 2). Although one of the strongest breast cancer risk factors (3, 4), not all women with benign breast disease (BBD) will develop breast cancer. To date, conventional approaches for risk stratification in BBD patients rely on microscopic assessment of epithelial abnormalities on BBD biopsies to classify women as having non-proliferative disease or proliferative disease without or with atypia (4-6). Patients with non-proliferative disease (∼70% of all BBD patients) are at minimal or no increased breast cancer risk (5). Proliferative diseases comprise ∼30% of all BBD biopsies and these patients have an almost 2-fold increased risk of breast cancer, with even higher 4-fold increased risk in the presence of atypical hyperplasia or multiple atypical foci (7). Notably, atypical hyperplasia diagnoses comprise only ∼4% of all BBD patients and, in absolute terms, fewer breast cancers will occur in these women than in those with non-proliferative BBD (8). Thus, there is the need to uncover additional tissue biomarkers that can aid to further stratify BBD patients into different breast cancer risk categories.

Microscopically, the normal breast is comprised of epithelial, stromal, and adipose tissue components (9). Although qualitative aberrations in epithelium underpin BBD-related breast cancer risk (10), the role of quantitative variation is poorly understood. Moreover, it remains fundamentally unclear whether risks related to BBD are driven by aberrations in the epithelium alone or via a dynamic interplay involving the stroma (11). Women undergoing breast biopsy, and for whom concomitant mammograms are available, represent an important patient population for the integrated study of histologic and radiologic breast tissue composition metrics in relation to breast cancer risk.

Within a cohort of women diagnosed as having BBD within a general community health care plan, we leveraged supervised machine-learning (12) and computer-assisted thresholding (13) methods to quantify breast tissue composition on histological and radiological images, respectively. This approach facilitated our investigations of the independent and joint associations of quantitative tissue metrics present at the time of BBD diagnosis with risk of subsequent breast cancer development.

## Methods and Materials

### Study population and design

We conducted a nested case-control study within a cohort of 15,395 women aged 18-86 years who were biopsied for BBD within the Kaiser Permanente Northwest Region (KPNW) between 1970-2012, with follow-up through mid-2015. KPNW is a prepaid healthcare plan with >500,000 members with facilities in Washington and Oregon. About 82% of KPNW members are White, 5% Asian American, 5% Hispanic, 3% African American and 5% other ethnicities (14). Case-control definition, ascertainment, and selection have been described in detail (15). Cases were women with a BBD biopsy who subsequently developed invasive breast cancer ≥one year after the index BBD biopsy. Controls were women biopsied for BBD who were alive but had not developed breast cancer during the same follow-up period as the corresponding case. Controls were selected using risk-set sampling and were individually matched to corresponding cases on age at BBD diagnosis (+/– 1 year) and plan membership duration. Data on breast cancer risk factors around the time of BBD diagnosis were manually abstracted from medical records (15).

### Acquisition and image analysis of BBD diagnostic hematoxylin and eosin (H&E)-stained sections

Diagnostic H&E slides were retrieved, and BBD lesions were classified according to Dupont and Page criteria as normal/non-proliferative, proliferative with atypia, and atypical hyperplasia (3). Terminal duct lobular unit (TDLU) involution was visually assessed based on published criteria from the Mayo BBD cohort (16) as follows: none (<25% of TDLUs involuted), partial (25-74%), or complete (≥75%) involution. Other histological information, including presence or absence of sclerosing adenosis, radial scar, ductal papilloma, fibroadenoma (simple or complex), and columnar cell change were obtained based on pathologists’ visual assessment (Yihong Wang and MAD).

H&E slides were scanned at high resolution (20×) using the Aperio digital slide scanner (Leica Biosystems Inc. Buffalo Grove, IL). Of the 1028 slides, 50 were unscannable due to quality control issues. Image analysis was performed using the Halo 1.2 Tissue Classifier algorithm (Indica Labs, Albuquerque, New Mexico). A 22-datapoint script involving two randomly selected representative-images was trained by a pathologist (MA) with expertise in digital pathology to identify, segment, and quantify (in mm^2^) areas on each slide comprised of epithelium (6-datapoints), stroma (5-datapoints), and adipose tissue (11-datapoints) as shown on **Figure 1 (A and B**). Training and centralized image analysis were performed masked to all patient characteristics. In reproducibility analysis, another pathologist (MAD) independently developed a 37-datapoint script to analyze a random sample of 185(∼20%) images. The results showed excellent agreement between the two pathologists’ scripts (Spearman’s rho=0.95, 0.97, and 0.98 for epithelium, stroma, and adipose tissue areas, respectively; **Supplementary Table 1**).

**Figure 1:**
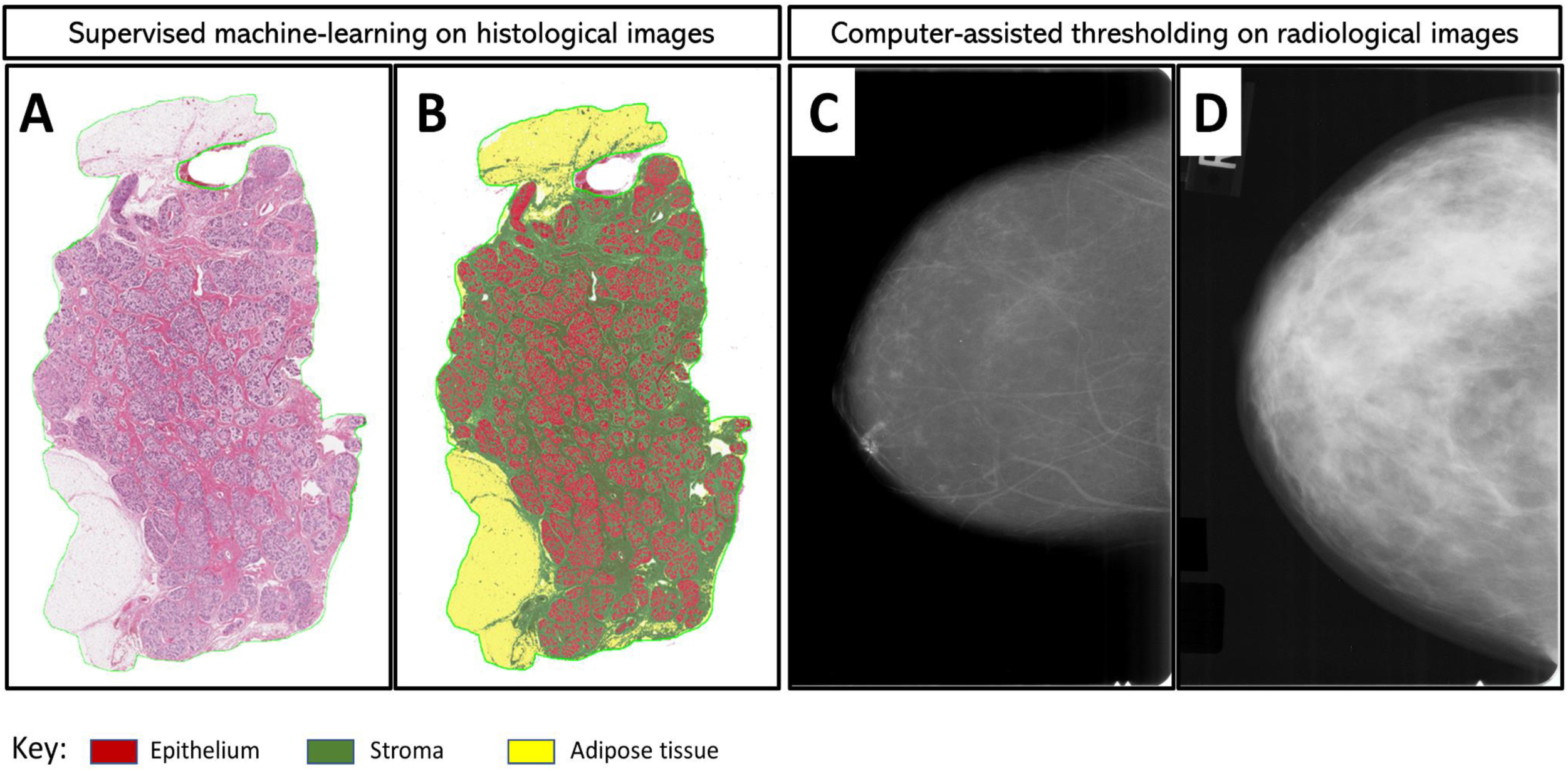
Quantitative assessment of breast tissue composition metrics from digitized histological and radiological images. Supervised machine-learning and computer-assisted thresholding methods were applied to histologic (A and B) and radiologic (C and D) images from women with BBD, respectively. Diagnostic hematoxylin and eosin (H&E) stained slides were digitized for image analysis while mammograms performed around the time of BBD diagnosis (average 1.3 months) were retrieved and digitized for analysis. For training purposes, a representative H&E image was randomly selected, and the machine was trained to identify areas of epithelium (red), stroma (green), and adipose tissue (yellow). Panel A is an example of an H&E image before analysis. In panel B, the machine learns-by-example to accurately classify and quantify epithelial (red), stromal (green), and adipose tissue (yellow) areas. Panels C and D are examples of representative mammograms that were determined to have low (below the median distribution among controls) and high (above the median) percent MBD based on quantitative assessment using the Cumulus software interface.

Percent epithelium, stroma, and adipose tissue were calculated by dividing the absolute value of each histologic metric by total tissue area on the slide and multiplying by 100. Given the documented biologic relevance of tumor-stroma ratio in the setting of cancer progression (17, 18), we sought to evaluate an equivalent feature in the context of BBD progression. Accordingly, we calculated the proportion of fibroglandular tissue (i.e. epithelium + stroma) on histology slides that was epithelium relative to stroma, i.e. histologic epithelium-to-stroma proportion (histologic-ESP), by dividing epithelial area by total fibroglandular tissue area and multiplying by 100.

### Mammogram retrieval and mammographic breast density assessment

The most recent mammograms occurring approximately 6 months before (preferably) or up to 1 month after the BBD biopsy were retrieved. Craniocaudal film mammographic views of the ipsilateral (preferable) or contralateral breast were digitized using an Array Corporation 2095 Laser Film Digitizer (Roden, the Netherlands; optical density=4.0). Quantitative measures of density were obtained using Cumulus®, an interactive computer-assisted thresholding program (19), with demonstrated validity with respect to breast cancer risk associations in numerous epidemiologic studies (20). All mammograms were evaluated by a single expert reader (EAB), who measured absolute dense area (cm^2^) and total breast area (cm^2^) as described previously (19). Percentage MBD was calculated by dividing the dense breast area by the total breast area and multiplying by 100 (**Figure 1 (C and D)**). Images from cases and matched controls were assessed within the same batch and in random order. A repeat set of 113 images was assessed for reliability. The intra-class correlation coefficients for percent MBD, dense area, and total breast area were 0.92, 0.89, and 0.99, respectively, documenting excellent reproducibility.

### Statistical analysis

Associations between baseline patient characteristics and tissue composition metrics were assessed in multivariable linear regression models fitted to controls. Locally weighted scatter plots of log residuals after regressing BMI and histology were used to demonstrate the distributions of tissue composition metrics by age among cases and controls. Quartiles (Q1-Q4) of tissue composition metrics were defined based on their distributions among controls. Associations between tissue composition metrics and breast cancer risk were assessed in crude and adjusted logistic regression models. For histologic metrics, conditional logistic regression models were adjusted for age at menarche, parity and age at first live birth, BMI, menopausal status/menopausal hormone therapy use, bilateral oophorectomy, history of breast cancer in a first degree relative, BBD histology, extent of lobular involution, calendar year of BBD diagnosis, as well as MBD. We used a likelihood ratio test to compare fit of a fully adjusted model with epithelium to a fully adjusted model with histologic-ESP. As radiologic tissue metrics were less complete for cases and controls, we used unconditional logistic regression, adjusting for matching factors (age at BBD diagnosis and follow-up duration), other risk factors noted above, as well as histologic-ESP, which showed better model fit than epithelium. To test the joint effects of histologic-ESP and MBD, both variables were dichotomized based on their median values among controls and a composite variable combining both was defined. Missing covariate values were imputed using the multiple (×5) imputation by chained equations (MICE) approach (21) with appropriate variance adjustment by Rubin’s Formula (22) for all analyses. Analyses were also stratified by BBD histological classification. P-values for heterogeneity were obtained by including multiplicative interaction terms between BBD histology and relevant risk factors in the full model. All analyses were two-sided and were performed using Stata statistical software version 16.1.

## Results

From 1,028 BBD patients, >95% (488 controls, 486 cases) had an H&E suitable for digitized pathology assessment, with a single image failing analysis. For radiologic metrics, a total of 302 (59%) controls and 297 (58%) cases had mammograms available within an average of 1.3 (standard deviation=3.5) months of BBD diagnosis. Most of the missing mammograms were for women diagnosed with BBD in the pre-screening (<1985) era. For those with BBD diagnosed in 1985 or thereafter, >85% of cases and controls had available mammograms for MBD assessment. In total, 564 patients (284 controls and 280 cases) had data on both histologic and radiologic metrics (**Supplementary Figure 1**). The median (range) age of patients at BBD diagnosis was 51.5 (18.7-86.6) years. BBD lesions were predominantly non-proliferative (69%), with fewer (28%) proliferative disease and atypical hyperplasia (3%). The distributions of other patient baseline characteristics are shown in **Table 1**.

**Table 1:**
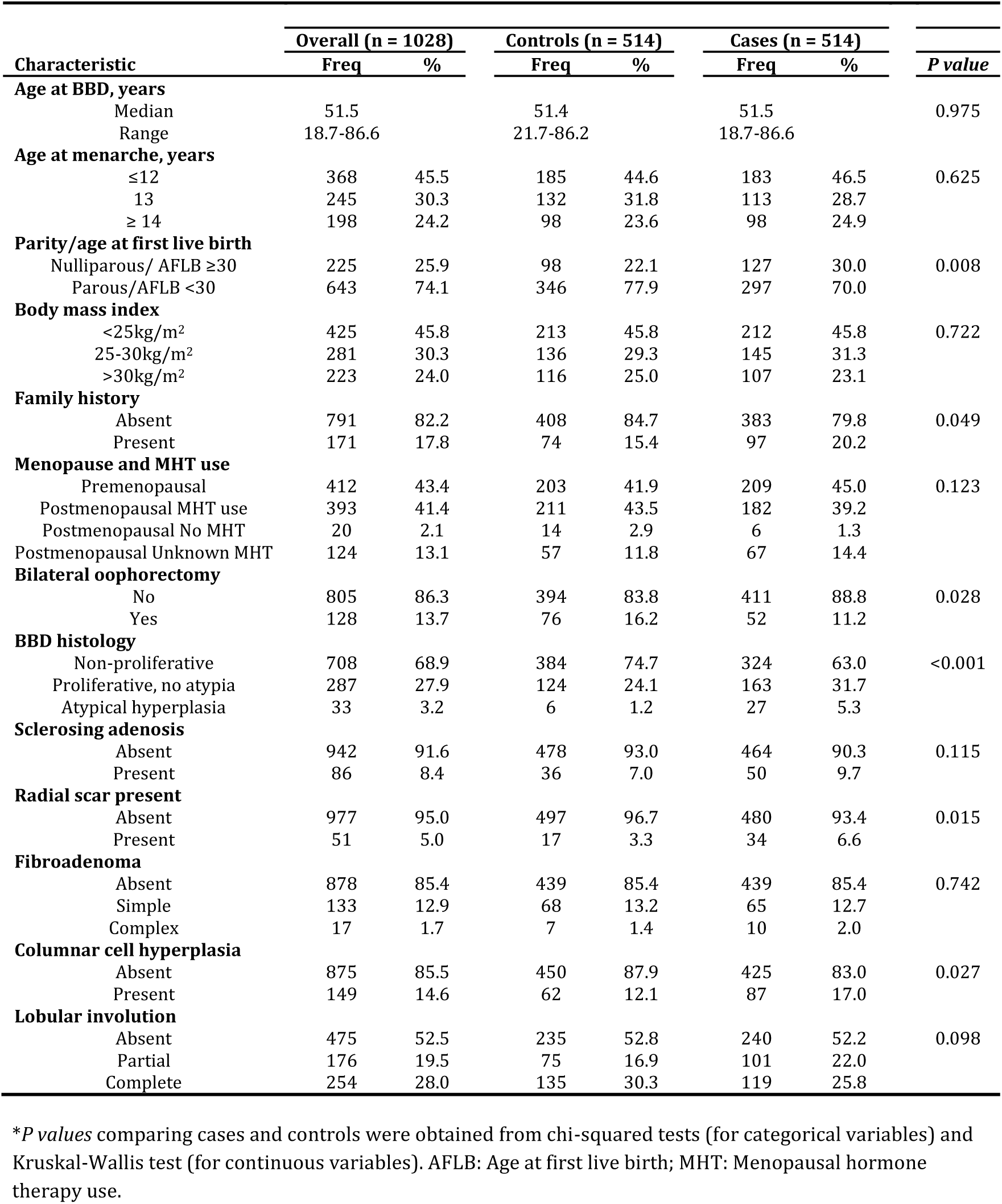
Baseline characteristics of the benign breast disease (BBD) patients, overall and by breast cancer case-control status, Kaiser Permanente Northwest Center for Health Research, 1970-2015

### Associations of histologic and radiologic breast tissue composition metrics with patients’ baseline characteristics

The median (range) of percent epithelial, stromal, adipose tissue and histologic-ESP distributions were 8.4% (0.2-97.4%), 38.1% (1.3-88.9%), 48.0% (1.3-97.5%), and 19.9% (0.9-98.6%), respectively. Medians (ranges) for absolute dense and non-dense areas and percent MBD, were 36.3cm^2^ (0-232.2cm^2^), 96.4cm^2^ (5.9-375.5cm^2^), and 30.2% (0-86.9%), respectively (**Supplementary Table 2**). The correlations between histologic and radiologic tissue composition metrics and their associations with baseline patient characteristics are provided in **Supplementary Tables 3 and 4**, respectively.

As shown in **Figure 2**, the fat component of the breast increased with advancing age whereas the fibroglandular tissue component declined, but rates of change differed between cases and controls. The rate of stromal change was fairly similar between cases and controls until around age 60 years after which it began to decline among cases, but not controls. Two peak differences were observed in histologic-ESP between cases and controls; first, between 40-50 years of age and second, from around 60-80 years of age.

**Figure 2:**
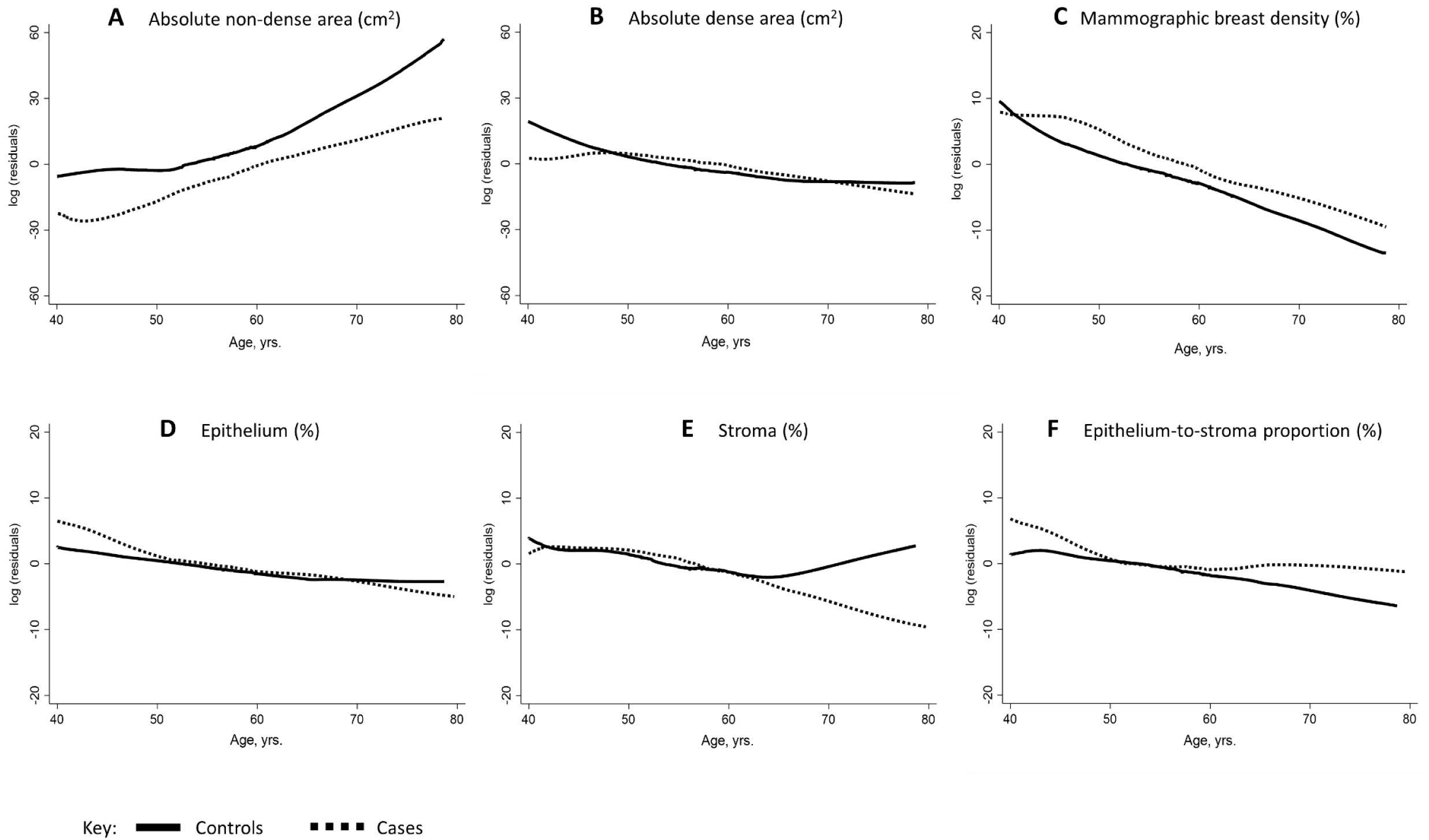
Histologic and radiologic breast tissue composition metrics by age and case-control status. Locally weighted scatter plot smoothing of log residuals (Y-axes) from linear regression models of non-dense area (A), dense area (B) and percent MBD (C), epithelium (D), stroma (E) and epithelium-to-stroma proportion (F). The effects of BMI and BBD histology on breast tissue composition were accounted for by adjusting for these in the linear regression models and subsequently plotting the log residuals against age.

### Associations between histologic tissue composition metrics and breast cancer risk

The median (range) time between BBD diagnosis and breast cancer incidence was 9 (1.5-38.5) years. As shown in **Table 2**, increasing epithelial content on BBD biopsies was dose-dependently associated with increasing breast cancer risk (OR(95% CI)_Q4 vs Q1_=1.85(1.13-3.04);P-trend=0.02), irrespective of BBD histology (P-heterogeneity=0.74). Conversely, the association between stroma and breast cancer risk differed by BBD histology (P-heterogeneity=0.002). Among women with non-proliferative disease, increasing stroma was associated with decreasing breast cancer risk (OR(95% CI)_Q4 vs Q1_=0.51(0.32-0.81);P-trend=0.006), whereas among those with proliferative disease it was associated with increasing risk (OR(95% CI)_Q4 vs Q1_=2.52(1.00, 6.32);P-trend=0.07). Histologic-ESP (LRχ^2^ =8.4; P=0.03) provided better model fit than epithelium (LRχ^2^ =7.4; P=0.06) and was associated with statistically significant increased risk of breast cancer (OR(95% CI)_Q4 vs Q1_=2.06(1.28, 3.33);P-trend=0.002), irrespective of BBD histology (P-heterogeneity=0.525). Notably, histologic-ESP remained statistically significantly associated with breast cancer risk even after adjusting for all other histologic features on diagnostic BBD histology slides as well as MBD (**Supplementary Table 5**).

**Table 2:** Odds ratios (ORs) and 95% confidence intervals (CIs) for the associations between histologic tissue composition metrics and risk of subsequent breast cancer development among women with BBD, overall and by BBD histological classification

| Histologic tissue metrics | Controls/Cases | Overall | Controls/Cases | Non-proliferative | Controls/Cases | Proliferative | *P* het |
| --- | --- | --- | --- | --- | --- | --- | --- |
|  |  | OR (95% CI) |  | OR (95% CI) |  | OR (95% CI) |  |
| **Epithelium area (%)** |  |  |  |  |  |  |  |
| <u>Quartiles</u> |  |  |  |  |  |  |  |
| Q1 (<4.45) | 122/103 | 1.00 (reference) | 100/83 | 1.00 (reference) | 22/20 | 1.00 (reference) | 0.741 |
| Q2 (4.45-7.93) | 122/111 | 1.10 (0.72, 1.67) | 100/78 | 1.02 (0.65, 1.62) | 22/33 | 1.30 (0.52, 3.27) |  |
| Q3 (7.93-14.36) | 122/111 | 1.13 (0.72, 1.76) | 90/64 | 0.97 (0.59, 1.57) | 32/47 | 1.54 (0.64, 3.74) |  |
| Q4 (>14.36) | 122/161 | 1.85 (1.13, 3.04) | 75/83 | 1.53 (0.93, 2.54) | 47/78 | 1.90 (0.81, 4.47) |  |
| *P* trend |  | 0.025 |  | 0.161 |  | 0.159 |  |
| Per 10% increase | 488/486 | 1.14 (0.99, 1.29) | 365/308 | 1.14 (0.98, 1.29) | 123/178 | 1.04 (0.86, 1.22) |  |
| *P* value |  | 0.066 |  | 0.087 |  | 0.647 |  |
| **Stroma area (%)** |  |  |  |  |  |  |  |
| <u>Quartiles</u> |  |  |  |  |  |  |  |
| Q1 (<24.66) | 122/134 | 1.00 (reference) | 83/94 | 1.00 (reference) | 39/40 | 1.00 (reference) | 0.002 |
| Q2 (24.66-38.08) | 122/107 | 0.62 (0.40, 0.95) | 85/59 | 0.48 (0.29, 0.79) | 37/48 | 1.09 (0.52, 2.27) |  |
| Q3 (38.08-53.70) | 122/124 | 0.67 (0.43, 1.05) | 87/68 | 0.48 (0.29, 0.79) | 35/56 | 1.23 (0.61, 2.51) |  |
| Q4 (>53.70) | 122/121 | 0.71 (0.46, 1.11) | 110/87 | 0.51 (0.32, 0.81) | 12/34 | 2.52 (1.00, 6.32) |  |
| *P* trend |  | 0.166 |  | 0.006 |  | 0.072 |  |
| Per 10% increase | 488/486 | 0.96 (0.87, 1.03) | 365/308 | 0.88 (0.80, 0.97) | 123/178 | 1.17 (1.01, 1.33) |  |
| *P* value |  | 0.248 |  | 0.010 |  | 0.037 |  |
| **Adipose tissue area (%)** |  |  |  |  |  |  |  |
| <u>Quartiles</u> |  |  |  |  |  |  |  |
| Q1 (<29.42) | 122/135 | 1.00 (reference) | 97/81 | 1.00 (reference) | 25/54 | 1.00 (reference) | 0.026 |
| Q2 (29.42-49.73) | 122/132 | 0.99 (0.65, 1.51) | 89/74 | 1.03 (0.65, 1.63) | 33/58 | 0.86 (0.42, 1.76) |  |
| Q3 (49.73-66.58) | 122/99 | 0.81 (0.54, 1.21) | 88/66 | 0.97 (0.61, 1.55) | 34/33 | 0.44 (0.20, 0.96) |  |
| Q4 (>66.58) | 122/120 | 1.15 (0.73, 1.81) | 91/87 | 1.61 (0.98, 2.64) | 31/33 | 0.56 (0.23, 1.34) |  |
| *P* trend |  | 0.884 |  | 0.094 |  | 0.065 |  |
| Per 10% increase | 488/486 | 1.01 (0.95, 1.08) | 365/308 | 1.06 (0.99, 1.14) | 123/178 | 0.93 (0.80, 1.06) |  |
| *P* value |  | 0.694 |  | 0.094 |  | 0.302 |  |
| **Histologic-ESP (%)** |  |  |  |  |  |  |  |
| <u>Quartiles</u> |  |  |  |  |  |  |  |
| Q1 (<10.82) | 122/93 | 1.00 (reference) | 109/97 | 1.00 (reference) | 13/16 | 1.00 (reference) | 0.525 |
| Q2 (10.82-18.23) | 122/110 | 1.24 (0.82, 1.88) | 99/79 | 1.20 (0.78, 1.87) | 23/31 | 1.08 (0.39, 2.95) |  |
| Q3 (18.24-29.01) | 122/131 | 1.58 (1.02, 2.45) | 85/72 | 1.40 (0.89, 2.23) | 37/59 | 1.41 (0.54, 3.66) |  |
| Q4 (>29.01) | 122/152 | 2.06 (1.28, 3.33) | 72/80 | 1.95 (1.21, 3.16) | 50/72 | 1.46 (0.57, 3.71) |  |
| *P* trend |  | 0.002 |  | 0.006 |  | 0.321 |  |
| Per 10% increase | 488/486 | 1.15 (1.04, 1.27) | 365/308 | 1.17 (1.06, 1.30) | 123/178 | 1.00 (0.85, 1.15) |  |
| *P* value |  | 0.011 |  | 0.004 |  | 0.979 |  |

Quartiles (Q1-Q4) of percent histologic tissue composition metrics (epithelium, stroma, adipose tissue, histologic epithelium-to-stroma proportion (histologic-ESP)) were defined based on their distributions among controls. In overall analyses, multivariate conditional logistic regression models adjusted for age at menarche, parity and age at first live birth, BMI, menopausal status/menopausal hormone therapy (MHT) use, bilateral oophorectomy, history of breast cancer in a first degree relative, BBD histology, extent of lobular involution, calendar year of BBD diagnosis, as well as MBD were used to estimate odds ratios (ORs) and corresponding 95% confidence intervals (CIs). In stratified analyses by BBD histology (i.e. non-proliferative disease and proliferative disease, with or without atypia), unconditional logistic regression models additionally adjusted for matching factors i.e. age at BBD diagnosis and follow-up time, were used to estimate ORs and 95% CIs. Epithelium and stroma were mutually adjusted for one another while histologic-ESP was additionally adjusted for adipose tissue. *P het*: P value for heterogeneity of OR estimates by BBD histological classification.

### Associations between radiologic tissue composition metrics and breast cancer risk

As shown in **Table 3**, increasing percent MBD was associated with increasing risk of breast cancer (OR(95% CI)_Q4 vs Q1_=2.20(1.20-4.03);P-trend=0.01), irrespective of BBD histology (P-heterogeneity=0.75).

**Table 3:**
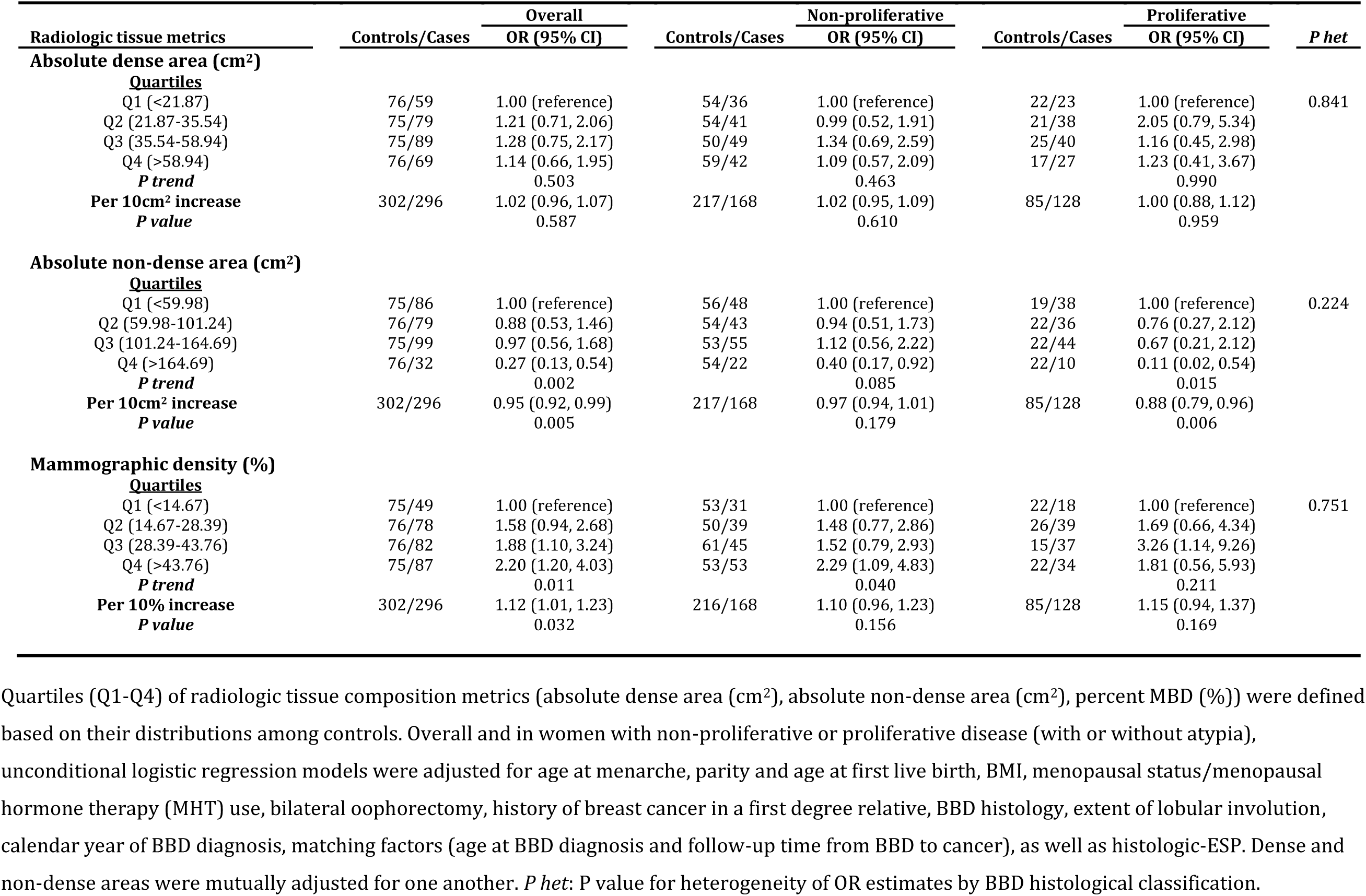
Odds ratios (ORs) and 95% confidence intervals (CIs) for the associations between radiologic tissue composition metrics and risk of subsequent breast cancer development among women with BBD, overall and by BBD histological classification

| Radiologic tissue metrics | Controls/Cases | Overall | Controls/Cases | Non-proliferative | Controls/Cases | Proliferative | *P* het |
| --- | --- | --- | --- | --- | --- | --- | --- |
|  |  | OR (95% CI) |  | OR (95% CI) |  | OR (95% CI) |  |
| Absolute dense area (cm<sup>2</sup>) |  |  |  |  |  |  |  |
| <u>Quartiles</u> |  |  |  |  |  |  |  |
| Q1 (<21.87) | 76/59 | 1.00 (reference) | 54/36 | 1.00 (reference) | 22/23 | 1.00 (reference) | 0.841 |
| Q2 (21.87-35.54) | 75/79 | 1.21 (0.71, 2.06) | 54/41 | 0.99 (0.52, 1.91) | 21/38 | 2.05 (0.79, 5.34) |  |
| Q3 (35.54-58.94) | 75/89 | 1.28 (0.75, 2.17) | 50/49 | 1.34 (0.69, 2.59) | 25/40 | 1.16 (0.45, 2.98) |  |
| Q4 (>58.94) | 76/69 | 1.14 (0.66, 1.95) | 59/42 | 1.09 (0.57, 2.09) | 17/27 | 1.23 (0.41, 3.67) |  |
| *P trend* |  | 0.503 |  | 0.463 |  | 0.990 |  |
| Per 10cm<sup>2</sup> increase | 302/296 | 1.02 (0.96, 1.07) | 217/168 | 1.02 (0.95, 1.09) | 85/128 | 1.00 (0.88, 1.12) |  |
| *P value* |  | 0.587 |  | 0.610 |  | 0.959 |  |
| Absolute non-dense area (cm<sup>2</sup>) |  |  |  |  |  |  |  |
| <u>Quartiles</u> |  |  |  |  |  |  |  |
| Q1 (<59.98) | 75/86 | 1.00 (reference) | 56/48 | 1.00 (reference) | 19/38 | 1.00 (reference) | 0.224 |
| Q2 (59.98-101.24) | 76/79 | 0.88 (0.53, 1.46) | 54/43 | 0.94 (0.51, 1.73) | 22/36 | 0.76 (0.27, 2.12) |  |
| Q3 (101.24-164.69) | 75/99 | 0.97 (0.56, 1.68) | 53/55 | 1.12 (0.56, 2.22) | 22/44 | 0.67 (0.21, 2.12) |  |
| Q4 (>164.69) | 76/32 | 0.27 (0.13, 0.54) | 54/22 | 0.40 (0.17, 0.92) | 22/10 | 0.11 (0.02, 0.54) |  |
| *P trend* |  | 0.002 |  | 0.085 |  | 0.015 |  |
| Per 10cm<sup>2</sup> increase | 302/296 | 0.95 (0.92, 0.99) | 217/168 | 0.97 (0.94, 1.01) | 85/128 | 0.88 (0.79, 0.96) |  |
| *P value* |  | 0.005 |  | 0.179 |  | 0.006 |  |
| Mammographic density (%) |  |  |  |  |  |  |  |
| <u>Quartiles</u> |  |  |  |  |  |  |  |
| Q1 (<14.67) | 75/49 | 1.00 (reference) | 53/31 | 1.00 (reference) | 22/18 | 1.00 (reference) | 0.751 |
| Q2 (14.67-28.39) | 76/78 | 1.58 (0.94, 2.68) | 50/39 | 1.48 (0.77, 2.86) | 26/39 | 1.69 (0.66, 4.34) |  |
| Q3 (28.39-43.76) | 76/82 | 1.88 (1.10, 3.24) | 61/45 | 1.52 (0.79, 2.93) | 15/37 | 3.26 (1.14, 9.26) |  |
| Q4 (>43.76) | 75/87 | 2.20 (1.20, 4.03) | 53/53 | 2.29 (1.09, 4.83) | 22/34 | 1.81 (0.56, 5.93) |  |
| *P trend* |  | 0.011 |  | 0.040 |  | 0.211 |  |
| Per 10% increase | 302/296 | 1.12 (1.01, 1.23) | 216/168 | 1.10 (0.96, 1.23) | 85/128 | 1.15 (0.94, 1.37) |  |
| *P value* |  | 0.032 |  | 0.156 |  | 0.169 |  |

Quartiles (Q1-Q4) of radiologic tissue composition metrics (absolute dense area (cm<sup>2</sup>), absolute non-dense area (cm<sup>2</sup>), percent MBD (%)) were defined based on their distributions among controls. Overall and in women with non-proliferative or proliferative disease (with or without atypia), unconditional logistic regression models were adjusted for age at menarche, parity and age at first live birth, BMI, menopausal status/menopausal hormone therapy (MHT) use, bilateral oophorectomy, history of breast cancer in a first degree relative, BBD histology, extent of lobular involution, calendar year of BBD diagnosis, matching factors (age at BBD diagnosis and follow-up time from BBD to cancer), as well as histologic-ESP. Dense and non-dense areas were mutually adjusted for one another. *P het*: P value for heterogeneity of OR estimates by BBD histological classification.

### Joint associations of histologic-ESP and MBD with breast cancer risk

Following dichotomization at their median values among controls, high histologic-ESP and percent MBD remained statistically significantly associated with elevated breast cancer risk (OR(95%CI)=1.57(1.13-2.18) and 1.50(1.01-2.24), respectively) (**Table 4)**. Further, patients with high histologic-ESP had higher breast cancer risk than those with low histologic-ESP, irrespective of whether they had high (OR(95%CI)=2.06(1.09-3.88) or low (OR(95%CI)=1.60(0.93-3.88) MBD (**Figure 3)**. Breast cancer risk was substantially higher in women with combined high histologic-ESP/high MBD than in those with low histologic-ESP/low MBD (OR(95%CI)=2.27(1.27-4.06);P-trend=0.005). These findings were stronger in patients with non-proliferative (2.43(1.20-4.93)) versus proliferative (1.55(0.45-5.33)) disease, though statistically significant heterogeneity was not observed (P-heterogeneity=0.73).

**Table 4:**
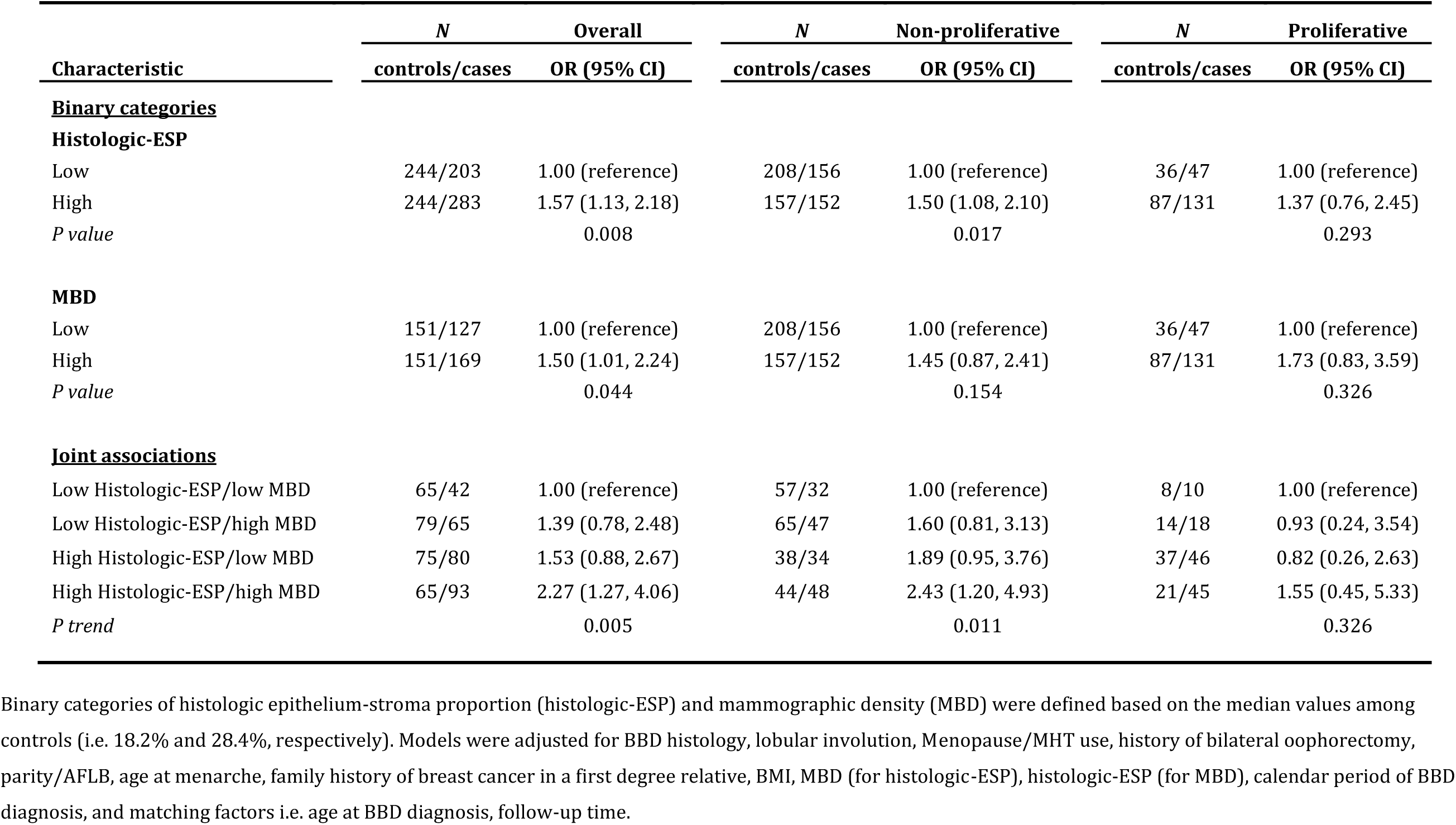
Odds ratios (ORs) and 95% confidence intervals (CIs) for the joint associations of epithelium-to-stroma proportion and percent MBD in relation to breast cancer risk among women with BBD, overall and by BBD histological classification.

| Characteristic | Overall |  | Non-proliferative |  | Proliferative |  |
| --- | --- | --- | --- | --- | --- | --- |
|  | *N*<br>controls/cases | OR (95% CI) | *N*<br>controls/cases | OR (95% CI) | *N*<br>controls/cases | OR (95% CI) |
| **<u>Binary categories</u>** |  |  |  |  |  |  |
| **Histologic-ESP** |  |  |  |  |  |  |
| Low | 244/203 | 1.00 (reference) | 208/156 | 1.00 (reference) | 36/47 | 1.00 (reference) |
| High | 244/283 | 1.57 (1.13, 2.18) | 157/152 | 1.50 (1.08, 2.10) | 87/131 | 1.37 (0.76, 2.45) |
| *P value* |  | 0.008 |  | 0.017 |  | 0.293 |
| **MBD** |  |  |  |  |  |  |
| Low | 151/127 | 1.00 (reference) | 208/156 | 1.00 (reference) | 36/47 | 1.00 (reference) |
| High | 151/169 | 1.50 (1.01, 2.24) | 157/152 | 1.45 (0.87, 2.41) | 87/131 | 1.73 (0.83, 3.59) |
| *P value* |  | 0.044 |  | 0.154 |  | 0.326 |
| **<u>Joint associations</u>** |  |  |  |  |  |  |
| Low Histologic-ESP/low MBD | 65/42 | 1.00 (reference) | 57/32 | 1.00 (reference) | 8/10 | 1.00 (reference) |
| Low Histologic-ESP/high MBD | 79/65 | 1.39 (0.78, 2.48) | 65/47 | 1.60 (0.81, 3.13) | 14/18 | 0.93 (0.24, 3.54) |
| High Histologic-ESP/low MBD | 75/80 | 1.53 (0.88, 2.67) | 38/34 | 1.89 (0.95, 3.76) | 37/46 | 0.82 (0.26, 2.63) |
| High Histologic-ESP/high MBD | 65/93 | 2.27 (1.27, 4.06) | 44/48 | 2.43 (1.20, 4.93) | 21/45 | 1.55 (0.45, 5.33) |
| *P trend* |  | 0.005 |  | 0.011 |  | 0.326 |

Binary categories of histologic epithelium-stroma proportion (histologic-ESP) and mammographic density (MBD) were defined based on the median values among controls (i.e. 18.2% and 28.4%, respectively). Models were adjusted for BBD histology, lobular involution, Menopause/MHT use, history of bilateral oophorectomy, parity/AFLB, age at menarche, family history of breast cancer in a first degree relative, BMI, MBD (for histologic-ESP), histologic-ESP (for MBD), calendar period of BBD diagnosis, and matching factors i.e. age at BBD diagnosis, follow-up time.

**Figure 3:**
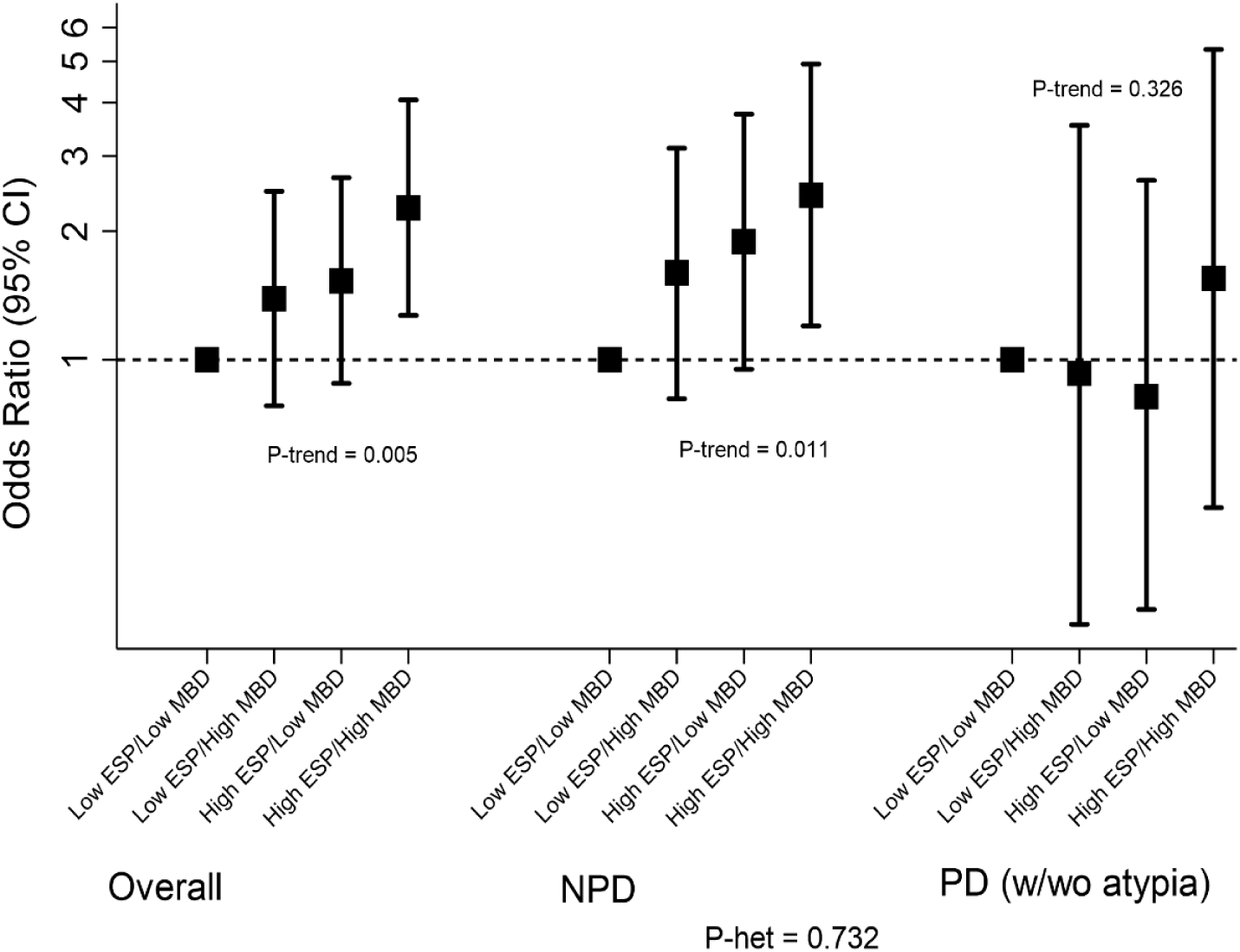
Joint associations of histologic-ESP and MBD and risk of subsequent breast cancer development among women with benign breast disease (BBD). Histologic-ESP and percent MBD were dichotomized at their median values among controls (i.e. 18.2% and 28.4%, respectively). Unconditional logistic regression models were adjusted for age at menarche, parity and age at first live birth, BMI, menopausal status/menopausal hormone therapy use, bilateral oophorectomy, history of breast cancer in a first degree relative, BBD histology, extent of lobular involution, calendar year of BBD diagnosis, as well as matching factors (age at BBD diagnosis and follow-up time from BBD to cancer). Analyses were performed overall (controls/cases (n) = 284/280) and among women with non-proliferative disease (NPD; controls/cases (n) = 204/161) and (C) proliferative disease (PD; controls/cases (n) = 80/119) BBD. Detailed odds ratios and related estimates are presented in **Table 4**.

In sensitivity analyses, both histologic-ESP and percent MBD were associated with elevated breast cancer risk, irrespective of menopausal status, calendar period of BBD diagnosis, or time between BBD diagnosis and breast cancer development. Further, histologic-ESP (OR(95% CI)_Q4 vs Q1_) more strongly predisposed to ER-positive (1.71(1.10-2.68);P-trend=0.009) than ER-negative (1.19(0.50-2.80);P-trend=0.37) as well as high-grade (2.08(1.25-3.44);P-trend=0.002) than low-grade (OR(1.25(0.65-2.39);P-trend=0.41) tumors, but differences by these tumor characteristics were not statistically significant.

## Discussion

To our knowledge, this is the first study to combine machine-learning and computer-assisted thresholding methods in the setting of BBD for histologic and radiologic assessments of tissue composition metrics, respectively, and to simultaneously relate these to breast cancer risk. We found dose-dependent relationships of histologic-ESP, a metric of the proportion of fibroglandular tissue on breast biopsies that is epithelium relative to stroma, and percent MBD, a metric of the proportion of total tissue area on mammograms that is radiodense, with risk of breast cancer development. Histologic-ESP and percent MBD were independently associated with risk; women with combined high histologic-ESP/high MBD had substantially higher breast cancer risk than those with low histologic-ESP/low MBD. The association between increasing stroma and breast cancer risk varied by the extent of epithelial hyperplasia; increasing stroma was associated with reduced risk in women with non-proliferative disease and increased risk in those with proliferative disease. This finding has not been reported previously and suggests a possible dual role of the stroma in mediating progression of breast precursor lesions. These results were robust to adjustments for other breast cancer risk factors. Taken together, our findings provide new insights into breast cancer development following BBD and could have implications for improved risk stratification and the clinical management of women with BBD, particularly those with non-proliferative disease, a large group for whom relevant predictive biomarkers are lacking.

To date, apart from BBD histological classification, very few risk factors for breast cancer have been identified for women with BBD. A few studies have reported the potential value of immunohistochemical (IHC) markers, including ER and/or PR expression, Ki-67 and CD20 in predicting risk, but these have yet to be consistently validated (23-26). Other reports support the value of TDLU involution in predicting breast cancer development following BBD (16, 27-29). However, these studies have largely been based on qualitative assessments of involution, with limited stratification. Although standardized measures of involution have been proposed (29), these are difficult to obtain and rely on the availability of “normal”, non-lesional tissue regions on BBD biopsies. Our findings of independent dose-response relationships of histologic-ESP and percent MBD with increasing breast cancer risk demonstrate the potential for these quantitative markers to improve risk stratification for BBD patients. Notably, histologic-ESP is a tissue-based feature that can easily be assessed on the same diagnostic H&E slides used for BBD diagnosis, without requiring IHC or other special stains. Accordingly, measures of histologic-ESP on BBD diagnostic H&E slides can be combined with MBD around the time of BBD diagnosis to provide additional information to women regarding their future breast cancer risk, at minimal or no extra cost or effort.

Despite experimental evidence to support a context-dependent role of stroma to either prevent or promote carcinogenesis (30-35), the precise sequence of events leading to a switch in stromal function from anti-to pro-tumorigenesis remains poorly understood. The prevailing model for BBD progression to cancer is that of a sequence of worsening epithelial abnormalities from normal/non-proliferative to proliferative disease (without atypia), atypical hyperplasia, in-situ carcinoma, and, ultimately, invasive breast cancer (36). Alternative pathways leading directly from normal/non-proliferative disease to invasive carcinoma have long been suspected (36), but specific tissue culprits are yet to be identified. Our finding of increasing breast cancer risk with increasing histologic-ESP that was particularly strong in women with normal/non-proliferative BBD supports an alternative model involving both epithelial and stromal compartments (**Figure 4**). In this model, the protective effect of the stroma is lost during the transition from non-proliferative to proliferative BBD and a “proliferative-switch” in stromal function from tumor-suppressor to tumor-promoter occurs.

**Figure 4:**
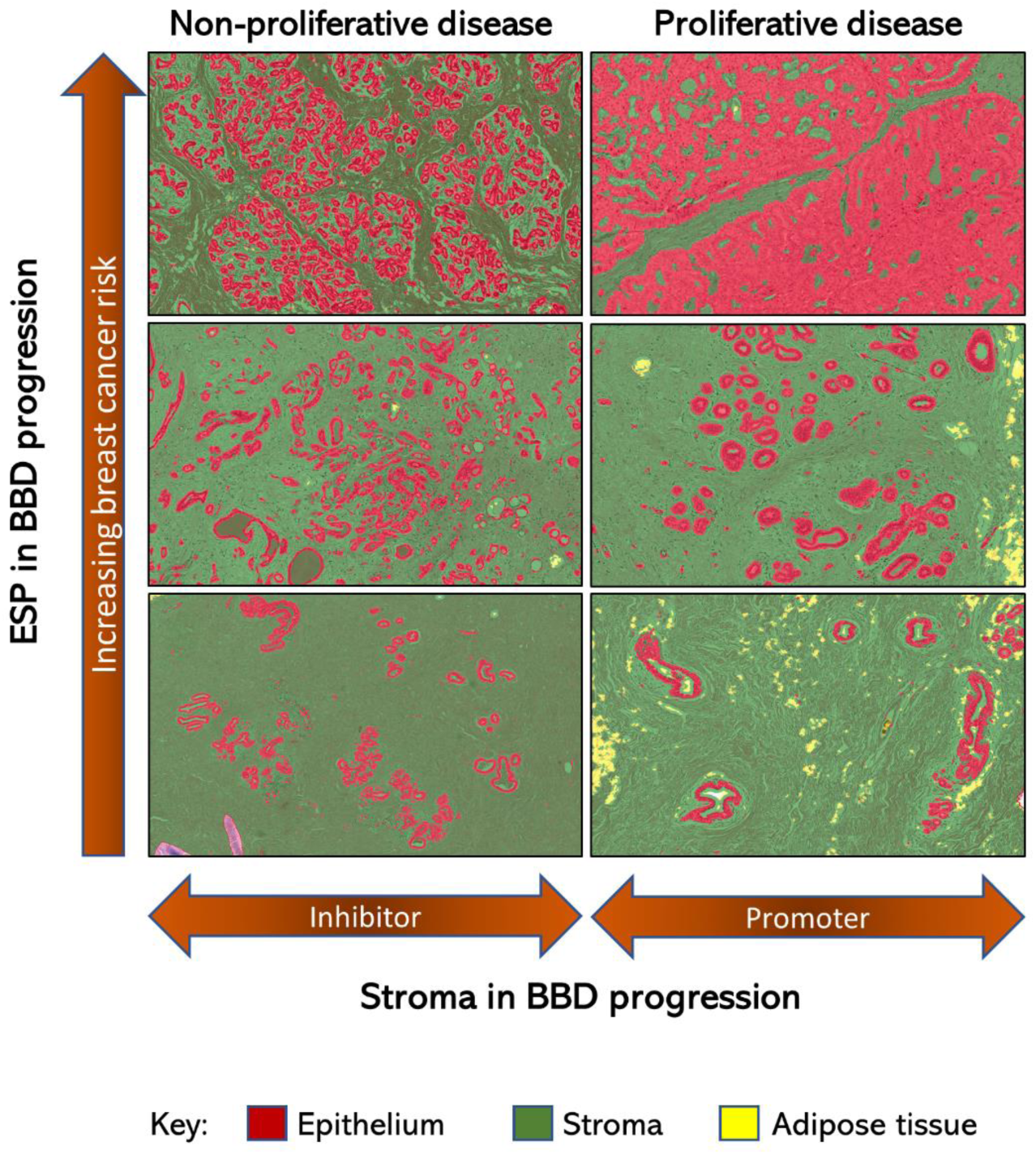
Conceptual model of BBD to breast cancer progression incorporating the contributions of changes in epithelium, stroma, and epithelium-to-stroma proportion to breast cancer risk. Increasing epithelium-to-stroma proportion (ESP) is displayed vertically, from bottom to top, to correspond to observed association with increasing risk of subsequent breast cancer development in this study (**Table 2**). The context-dependent role of the stroma to either inhibit or promote tumor formation in the setting of non-proliferative or proliferative disease (**Table 2**), respectively, is displayed horizontally. In this conceptual model of BBD to breast cancer progression, we propose that the proportion of the epithelial and stromal components of the breast is in a delicate balance during normal homeostasis. Disruption of this balance, either through uncontrolled epithelial proliferation, lack of age-related epithelial involution, or stromal depletion, will manifest as increasing ESP (**Supplementary Table 4**). High ESP may, in turn, represent a feature of the breast microenvironment that is conducive to carcinogenesis.

An important aspect in the clinical management of women with BBD or high MBD is to decide who is at sufficiently high risk to benefit from preventative strategies, such as chemoprevention, that reduce risk of developing cancer. Available risk prediction tools (37-42) have modest discriminatory accuracy, which could be improved by adding quantitative tissue composition metrics such as histologic-ESP and percent MBD. Furthermore, as >43% of screened U.S. women have dense breasts (43), it is imperative to identify additional factors that may identify those at high risk of invasive disease, requiring further clinical management (44, 45). In the current study, high histologic-ESP portended elevated breast cancer risk for women with low *or* high MBD, buttressing the importance of integrating histologic measures, when available, for distinguishing relative proportions of epithelium and stroma in radiodense tissue, a distinction that cannot currently be made on mammograms.

Our application of machine-learning to digitized H&E slides allowed us to perform centralized analysis of all images using a single script, thereby limiting subject-specific bias and random error. The correlation between different scripts that were independently trained by two pathologists was excellent. This study is, however, not without limitations: most patients in this analysis underwent excisional biopsies, which have been largely replaced by needle biopsies as the standard of care. Also, there were too few women with atypical hyperplasia to allow for separate analysis. Future work integrating more contemporary approaches, including artificial intelligence (46), for density assessment on digital mammography will be important for extending findings.

In summary, quantitative assessments of histologic-ESP on diagnostic BBD biopsy slides and percent MBD on mammograms performed around the time of BBD diagnosis demonstrated dose-dependent relationships with increasing risk of subsequent invasive breast cancer development, particularly for women with non-proliferative disease. Furthermore, histologic-ESP identified women with low MBD who were at significantly elevated risk of breast cancer and those with high MBD who were not. We also uncovered a context-dependent role of the stroma to either decrease or increase breast cancer risk in women with non-proliferative versus proliferative disease, respectively. Taken together, these findings provide clues regarding breast cancer etiology and BBD progression and could have implications for risk stratification and the clinical management of women with benign findings upon breast biopsy.

## Data Availability

The data underlying this article will be shared on reasonable request to the corresponding author.

## Funding

This work was supported by Intramural Research Funds of the Division of Cancer Epidemiology and Genetics (DCEG) of the National Cancer Institute, Department of Health and Human Services, USA.

Thomas Rohan is supported by the Breast Cancer Research Foundation (BCRF-19-140). Erin Bowles is supported by the National Cancer Institute R50CA211115.

## Notes

### Funder’s Role

The funder had no role in the design of the study; the collection, analysis, and interpretation of the data; the writing of the manuscript; and the decision to submit the manuscript for publication. The authors have no disclosures

### Data availability statement

The data underlying this article will be shared on reasonable request to the corresponding author.

## Acknowledgements

We thank Dr. Yihong Wang who performed the BBD pathology evaluations.

## Conflicts of interest

The authors declare no conflicts of interest.

## Author contributions

Conception and design: MA, AGG, JDF, TER, GLG

Provision of study materials or patients: KR, AGG, TMK, TER

Generation, collection, and assembly of data: MA, SF, EAB, LW, MAD, RMP, RTF, SL, KR, AGG, TMK, TER

Data analysis and interpretation: All authors

Administrative support: RTF, SL, TMK

Manuscript writing: MA, SF, MAD, RMP, JDF, GLG

Final approval of manuscript: All authors have seen and approved the manuscript.

## Supplementary material

### Tables

**Supplementary Table 1**: Reproducibility analysis between pathologists’ machine-learning scripts for tissue composition analysis on H&E-stained slides of BBD biopsies

**Supplementary Table 2**: Distributions of histologic and radiologic breast tissue composition metrics in women with benign breast disease, overall and among cases and controls

**Supplementary Table 3**: Correlations between quantitative histologic and radiologic breast tissue composition metrics obtained around the time of BBD diagnosis for women with BBD

**Supplementary Table 4**: Association between baseline patient characteristics and tissue composition metrics among controls

**Supplementary Table 5**: Odds ratios (ORs) and 95% confidence intervals (CIs) for the associations between individual histologic features on BBD biopsy and risk of subsequent breast cancer development among women with BBD

### Figures

**Supplementary Figure 1**: Study population.

## Notes

### Competing Interest Statement

The authors have declared no competing interest.

### Author Declarations

NIH/NCI/DCEG Institutional Review Board

