## Supplementary Tables for "Relation of Quantitative Histologic and Radiologic Breast Tissue Composition Metrics with Invasive Breast Cancer Risk"

**Supplementary Figure 1:** Study population

**Supplementary Table 1**: Reproducibility analysis between pathologists' machine-learning scripts for tissue composition analysis on H&E-stained slides of BBD biopsies

**Supplementary Table 2**: Distributions of histologic and radiologic breast tissue composition metrics in women with benign breast disease, overall and among cases and controls

**Supplementary Table 3**: Correlations between quantitative histologic and radiologic breast tissue composition metrics obtained around the time of BBD diagnosis for women with BBD

**Supplementary Table 4**: Association between baseline patient characteristics and tissue composition metrics among controls

**Supplementary Table 5**: Odds ratios (ORs) and 95% confidence intervals (CIs) for the associations between individual histologic features on BBD biopsy and risk of subsequent breast cancer development among women with BBD

**Supplementary Figure 1**: Study population

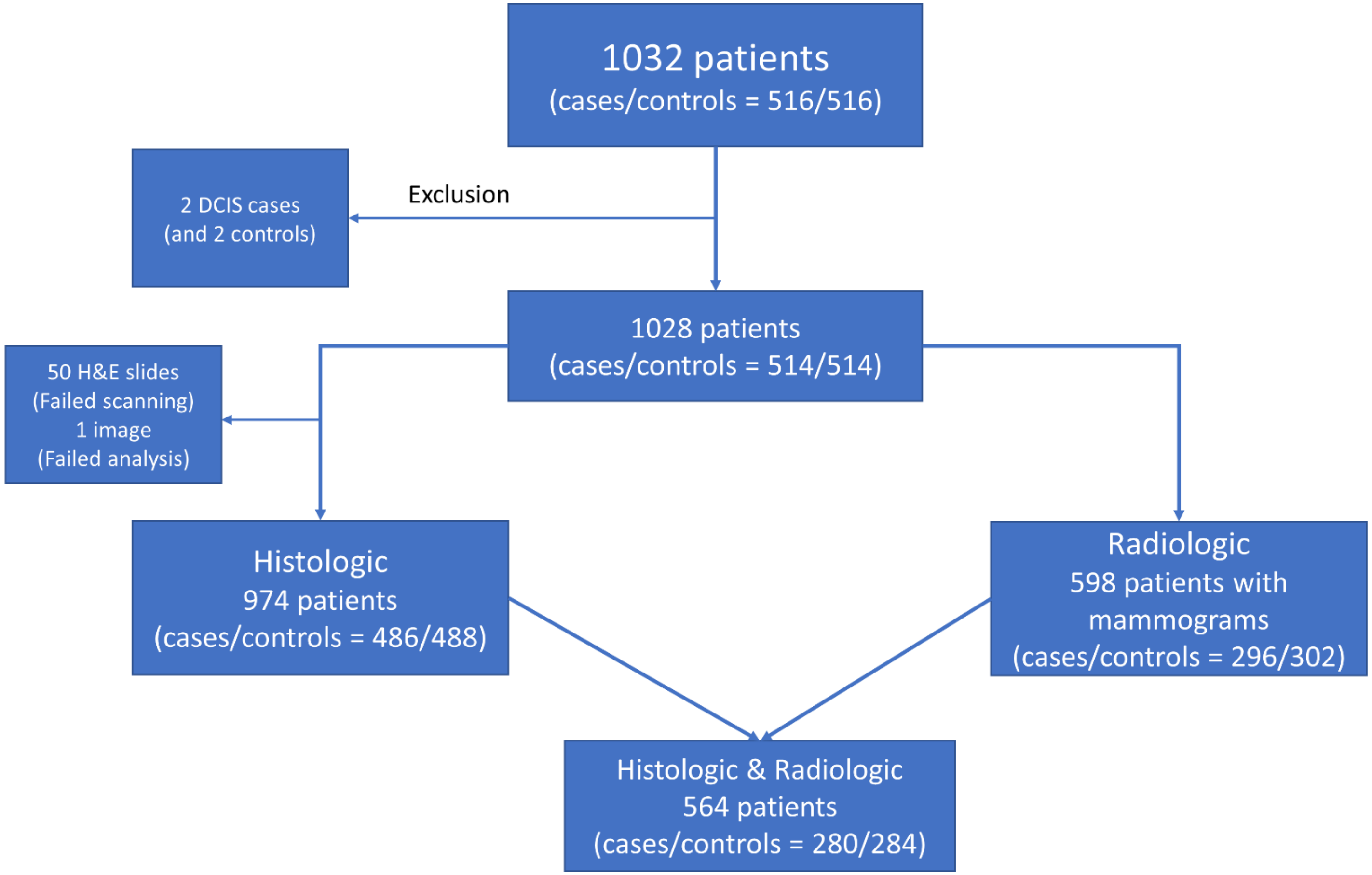

**Supplementary Table 1**: Reproducibility analysis between two pathologists' machine-learning scripts for tissue composition analysis on digitized H&E-stained slides

A) Spearman correlation coefficients for the agreement between deciles of histologic tissue composition features derived using two independently trained scripts

| | | Pathologist 1 | | |
|---|---|---|---|---|
| | | Epithelium | Stroma | Adipose tissue |
| Pathologist 2 | **Epithelium** | 0.95 | | |
| | ***P value*** | <0.001 | | |
| | **Stroma** | | 0.97 | |
| | ***P value*** | | <0.001 | |
| | **Adipose tissue** | | | 0.98 |
| | ***P value*** | | | <0.001 |

B) Inter-rater agreement between deciles of histologic tissue composition features derived using two independently trained scripts

| | | Pathologist 1 | | | | | |
|---|---|---|---|---|---|---|---|
| | | Epithelium | | Stroma | | Adipose tissue | |
| | | Agreement | Kappa | Agreement | Kappa | Agreement | Kappa |
| Pathologist 2 | **Epithelium** | 94% | 0.84 | | | | |
| | ***P value*** | <0.0001 | | | | | |
| | **Stroma** | | | 96% | 0.90 | | |
| | ***P value*** | | | <0.0001 | | | |
| | **Adipose tissue** | | | | | 96% | 0.90 |
| | ***P value*** | | | | | <0.0001 | |

**Supplementary Table 2:** Distributions of histologic and radiologic breast tissue composition metrics in women with benign breast disease, overall and among cases and controls

| | Controls | | | | Cases | | | | |
|---|---|---|---|---|---|---|---|---|---|
| **Characteristic** | ***n*** | **Mean** | **Median** | **Range** | ***n*** | **Mean** | **Median** | **Range** | ***P*** |
| **Histological** | | | | | | | | | |
| % Epithelium | 488 | 11.8 | 7.9 | 0.2-97.4 | 486 | 13.5 | 8.9 | 0.6-75.6 | 0.096 |
| % Stroma | 488 | 39.6 | 38.1 | 1.3-88.9 | 486 | 39.0 | 38.2 | 1.8-86.2 | 0.898 |
| % Adipose tissue | 488 | 48.6 | 49.7 | 1.3-95.7 | 486 | 47.4 | 46.6 | 3.0-97.5 | 0.201 |
| % Histologic-ESP | 488 | 22.4 | 18.2 | 0.9-98.6 | 486 | 24.9 | 21.4 | 0.9-88.6 | 0.021 |
| **Radiological** | | | | | | | | | |
| Absolute dense area ($cm^2$) | 302 | 44.1 | 35.5 | 0.0-232.2 | 296 | 44.9 | 36.6 | 3.7-208.1 | 0.513 |
| Absolute non-dense area ($cm^2$) | 302 | 117.7 | 101.2 | 5.9-368.6 | 296 | 101.5 | 86.5 | 8.0-375.5 | 0.141 |
| % Mammographic density | 302 | 31.2 | 28.4 | 0.0-84.01 | 296 | 34.2 | 31.9 | 0.9-86.9 | 0.102 |

Histologic-ESP: Histologic epithelium-to-stroma proportion

**Supplementary Table 3**: Correlations between quantitative histologic and radiologic breast tissue composition metrics obtained around the time of BBD diagnosis for women with BBD

| | | Histologic | | | | Radiologic | | |
|---|---|---|---|---|---|---|---|---|
| | | **Epithelium** | **Stroma** | **Adipose tissue** | **Histologic-ESP** | **Dense area** | **Non-dense area** | **MBD** |
| | Dense area | -0.005 | 0.202 | -0.165 | -0.131 | 1.000 | | |
| | *P value* | 0.911 | <0.001 | <0.001 | 0.002 | | | |
| **Radiologic** | Non-dense area | -0.012 | -0.272 | 0.279 | 0.011 | -0.089 | 1.000 | |
| | *P value* | 0.015 | <0.001 | <0.001 | 0.787 | 0.030 | | |
| | MBD | 0.082 | 0.338 | -0.323 | -0.074 | 0.619 | -0.691 | 1.000 |
| | *P value* | 0.051 | <0.001 | <0.001 | 0.081 | <0.001 | <0.001 | |
| | Epithelium | 1.000 | | | | | | |
| | *P value* | | | | | | | |
| | Stroma | 0.014 | 1.000 | | | | | |
| **Histologic** | *P value* | 0.667 | | | | | | |
| | Adipose tissue | -0.556 | -0.838 | 1.000 | | | | |
| | *P value* | <0.001 | <0.001 | | | | | |
| | Histologic-ESP | 0.824 | -0.416 | -0.104 | 1.000 | | | |
| | *P value* | <0.001 | <0.001 | 0.001 | | | | |

Histologic-ESP: histologic epithelium-to-stroma proportion; MBD: Mammographic breast density; N = 564 women had information on both histologic and radiologic breast tissue composition metrics.

**Supplementary Table 4**: Associations between baseline patient characteristics and tissue composition metrics among controls

| | Histologic tissue composition (N = 976) | | | | | | | | Radiologic tissue composition[ǂ] (N = 601) | | | | | |
|---|---|---|---|---|---|---|---|---|---|---|---|---|---|---|
| | Epithelium[ǂ] | | Stroma | | Adipose | | histologic-ESP[ǂ] | | Dense area | | Non-dense area | | MBD[Δ] | |
| **Characteristic** | **β** | **p** | **β** | **p** | **β** | **p** | **β** | **p** | **β** | **p** | **β** | **p** | **β** | **p** |
| **Age at BBD, yrs.** | -0.20 | **<0.001** | 0.04 | 0.971 | 0.23 | 0.096 | -0.14 | **<0.001** | -0.05 | 0.475 | 0.09 | **0.045** | -0.01 | 0.130 |
| **BBD histology[φ]** | | | | | | | | | | | | | | |
| PD | 0.53 | **<0.001** | -7.69 | **0.002** | 0.39 | 0.894 | 0.52 | **<0.001** | -0.09 | 0.547 | -0.03 | 0.757 | -0.07 | 0.603 |
| AH | 0.32 | 0.331 | -5.55 | 0.470 | 1.76 | 0.844 | 0.32 | 0.238 | 0.13 | 0.748 | 0.07 | 0.804 | 0.03 | 0.944 |
| P trend | | **<0.001** | | **0.005** | | 0.932 | | **<0.001** | | 0.656 | | 0.978 | | 0.592 |
| **Sclerosing adenosis present** | 0.15 | 0.364 | -2.69 | 0.494 | -0.63 | 0.890 | 0.16 | 0.258 | 0.30 | 0.199 | 0.03 | 0.827 | 0.20 | 0.349 |
| **Radial scar present** | 0.23 | 0.314 | 0.44 | 0.935 | -3.20 | 0.606 | 0.14 | 0.466 | -0.32 | 0.285 | 0.16 | 0.419 | -0.05 | 0.861 |
| **Simple fibroadenoma** | 0.55 | **<0.001** | 7.96 | **0.002** | -14.48 | **<0.001** | 0.23 | **0.013** | -0.47 | **0.003** | 0.02 | 0.879 | -0.40 | **0.008** |
| **Complex fibroadenoma** | 0.38 | 0.302 | 3.52 | 0.687 | -3.20 | 0.606 | 0.19 | 0.533 | -0.42 | 0.320 | -0.02 | 0.949 | -0.22 | 0.581 |
| **Columnar cell hyperplasia** | 0.18 | 0.124 | 6.13 | **0.024** | -6.83 | **0.031** | -0.01 | 0.968 | 0.15 | 0.391 | -0.18 | 0.125 | 0.24 | 0.148 |
| **BMI** | | | | | | | | | | | | | | |
| 25-30kg/m2 | -0.11 | 0.331 | 2.12 | 0.355 | -0.11 | 0.968 | -0.12 | 0.164 | -0.21 | 0.103 | 0.59 | **<0.001** | -0.57 | **<0.001** |
| >30kg/m2 | -0.16 | 0.254 | -2.30 | 0.402 | 4.13 | 0.209 | -0.06 | 0.577 | -0.08 | 0.539 | 0.98 | **<0.001** | -0.76 | **<0.001** |
| P trend | | 0.244 | | 0.463 | | 0.219 | | 0.495 | | 0.470 | | **<0.001** | | **<0.001** |
| **Positive family history** | 0.16 | 0.142 | -5.15 | **0.037** | 2.29 | 0.416 | 0.21 | **0.014** | 0.12 | 0.391 | 0.01 | 0.893 | 0.09 | 0.476 |
| **Bilateral oophorectomy** | -0.27 | **0.029** | -5.22 | **0.040** | 7.62 | **0.011** | -0.03 | 0.799 | -0.08 | 0.588 | 0.10 | 0.276 | -0.12 | 0.402 |
| **Postmenopausal MHT use** | 0.11 | 0.350 | 0.07 | 0.979 | -1.62 | 0.606 | 0.05 | 0.589 | -0.08 | 0.616 | -0.13 | 0.233 | 0.04 | 0.781 |
| **Postmenopausal No MHT** | 0.19 | 0.409 | -10.80 | 0.065 | 10.86 | 0.105 | 0.42 | 0.050 | -0.30 | 0.326 | 0.17 | 0.385 | -0.31 | 0.289 |
| **Parity and age at first live birth[¥]** | | | | | | | | | | | | | | |
| Nulliparous/ AFLB ≥30 | | | | | | | | | | | | | | |
| Parous/AFLB <30 | 0.07 | 0.468 | -2.21 | 0.295 | 1.85 | 0.472 | 0.13 | 0.096 | -0.28 | **0.026** | 0.22 | **0.018** | -0.33 | **0.007** |
| **Menarche** | | | | | | | | | | | | | | |
| 12 years (*Ref*) | | | | | | | | | | | | | | |
| 13 years | -0.04 | 0.682 | -1.26 | 0.581 | 1.05 | 0.679 | -0.01 | 0.971 | 0.05 | 0.707 | 0.15 | 0.899 | 0.05 | 0.730 |
| ≥ 14 years | -0.07 | 0.438 | -1.81 | 0.446 | 1.66 | 0.541 | -0.07 | 0.337 | 0.01 | 0.917 | 0.06 | 0.454 | -0.04 | 0.779 |
| P trend | | 0.367 | | 0.502 | | 0.560 | | 0.282 | | 0.754 | | 0.626 | | 0.970 |
| **Lobular involution** | | | | | | | | | | | | | | |
| Partial | -0.33 | **0.006** | 2.26 | 0.396 | 1.04 | 0.709 | -0.28 | **0.021** | 0.23 | 0.121 | -0.18 | 0.069 | 0.29 | **0.042** |
| Complete | -0.53 | **<0.001** | -2.78 | 0.212 | 6.95 | **0.010** | -0.31 | **<0.001** | 0.17 | 0.238 | -0.06 | 0.498 | 0.17 | 0.241 |
| P trend | | **<0.001** | | 0.200 | | **0.010** | | **0.001** | | 0.267 | | 0.481 | | 0.265 |
| **% MBD[₮]** | | | | | | | | | | | | | | |
| Q1 (*Ref*) | | | | | | | | | | | | | | |
| Q2 | -0.15 | 0.245 | 6.34 | 0.074 | -3.75 | 0.372 | -0.24 | **0.039** | | | | | | |
| Q3 | -0.02 | 0.878 | 4.73 | 0.151 | -3.43 | 0.390 | -0.11 | 0.336 | | | | | | |
| Q4 | -0.08 | 0.578 | 10.92 | **0.008** | -9.54 | **0.029** | -0.30 | **0.011** | | | | | | |
| P trend | | 0.772 | | **0.005** | | **0.022** | | **0.030** | | | | | | |

[ǂ] Log transformed to approximate normality. [φ]PD = Proliferative disease, no atypia; AH = Atypical hyperplasia. [¥]AFLB = Age at first live birth. [§]Menopausal hormone therapy. Histologic-ESP = epithelium-to-stroma proportion; MBD = mammographic breast density; [Δ]MBD continuous measure; [₮]MBD quartiles based on distribution in controls.

**Supplementary Table 5**: Odds ratios (ORs) and 95% confidence intervals (CIs) for the associations between individual histologic features on BBD biopsy and risk of subsequent breast cancer development among women with BBD

| Characteristic | controls/cases | OR (95% CI) |
|---|---|---|
| **Histologic-ESP** | | |
| Q1 (<10.82) | 122/93 | 1.00 (reference) |
| Q2 (10.82-18.24) | 122/110 | 1.34 (0.89, 2.00) |
| Q3 (18.24-29.13) | 122/131 | 1.57 (1.03, 2.41) |
| Q4 (>29.13) | 122/152 | 2.10 (1.33, 3.32) |
| *P trend* | | 0.002 |
| Per 10% increase | 488/486 | 1.15 (1.04, 1.26) |
| *P value* | | 0.008 |
| **Sclerosing adenosis** | | |
| Absent | 454/440 | 1.00 (reference) |
| Present | 34/46 | 0.99 (0.59, 1.68) |
| **Radial Scar** | | |
| Absent | 472/456 | 1.00 (reference) |
| Present | 16/30 | 1.36 (0.66, 2.81) |
| **Fibroadenoma** | | |
| Absent | 417/412 | 1.00 (reference) |
| Simple | 66/64 | 1.01 (0.66, 1.64) |
| Complex | 5/10 | 2.39 (0.71, 7.97) |
| **Columnar cell change** | | |
| None | 429/402 | 1.00 (reference) |
| Present | 57/83 | 1.74 (1.11, 2.72) |

Quartiles (Q1-Q4) of histologic epithelium-stroma proportion (histologic-ESP) were defined based on the distribution among controls. Models were additionally adjusted for lobular involution, BMI, parity/AFLB, oophorectomy, calendar period of BBD diagnosis, matching factors i.e. age at BBD diagnosis, follow-up time, as well as MBD.
